# The effect of stimulus duration on preferences for gain adjustments when listening to speech

**DOI:** 10.1101/2021.02.26.21252511

**Authors:** William M. Whitmer, Benjamin Caswell-Midwinter, Graham Naylor

**Affiliations:** Hearing Sciences - Scottish Section, School of Medicine, University of Nottingham, Glasgow, UK; Institute of Health and Wellbeing, College of Medical, Veterinary and Life Science, University of Glasgow, Glasgow, UK; Otolaryngology - Head and Neck Surgery, Massachusetts Eye and Ear, Harvard Medical School, Boston, Massachusetts, USA

## Abstract

**Objectives:** In the personalisation of hearing aid fittings, gain is often adjusted to suit patient preferences using live speech. When using brief sentences as stimuli, the minimum gain adjustments necessary to elicit consistent preferences (‘preference thresholds’) were previously found to be much greater than typical adjustments in current practice. The current study examined the role of duration on preference thresholds.

**Design:** Participants heard 2, 4 and 6-s segments of a continuous monologue presented successively in pairs. The first segment of each pair was presented at each individual’s real-ear or prescribed gain. The second segment was presented with a ±0-12 dB gain adjustment in one of three frequency bands. Participants judged whether the second was “better”, “worse” or “no different” from the first.

**Study Sample:** Twenty-nine adults, all with hearing-aid experience.

**Results:** The minimum gain adjustments needed to elicit “better” or “worse” judgments decreased with increasing duration for most adjustments. Inter-participant agreement and intra-participant reliability increased with increasing duration up to 4 s, then remained stable.

**Conclusions:** Providing longer stimuli improves the likelihood of patients providing reliable judgments of hearing-aid gain adjustments, but the effect is limited, and alternative fitting methods may be more viable for effective hearing-aid personalisation.

## Introduction

In the treatment of hearing loss, clinicians fit hearing aids to reach a balance between audibility and comfort for each patient. The balancing begins with prescribed gains across frequency based on each patient’s pure-tone thresholds. These prescribed gains, based on average data, are then personalised through adjustments made by the clinician using patient feedback (Anderson et al., 2018; Jenstad et al., 2003; Kuk, 1999; Thielemans et al., 2017). The patient’s feedback is often based solely on the effect the adjustments have on the perception of the clinician’s voice, the most readily available stimulus in any clinic.

We have previously investigated what gain adjustments are discriminable for short sentences presented in quiet. Median just-noticeable differences (JNDs) for increases in gain (increments) in broad low-, mid- and high-frequency bands were 4, 4 and 7 dB, respectively (Caswell-Midwinter and Whitmer, 2019). Gain adjustments less than these JNDs will, on average, not be readily perceived. A clinician may still receive feedback from a patient, but such feedback may not be based on the auditory perception of these adjustments, but other factors (cf. placebo effects without adjustment; Bentler et al., 2003; Dawes et al., 2013; Naylor et al., 2015). Using the same speech corpus, we have subsequently investigated what gain adjustments are necessary to elicit consistent preferences (Caswell-Midwinter and Whitmer, 2020). Median preference thresholds, the minimum adjustment to elicit a preference, ranged from 4-12 dB for gain decrements and 5-9 dB for increments in the same broad low-, mid-, and high-frequency bands. In Caswell-Midwinter and Whitmer (2019), it was posited that the greater JNDs for speech in quiet than for speech-shaped noise were due to the spectro-temporal sparsity of the speech. That is, for a given gain adjustment in any given band, the clean speech signal provided a smaller number of glimpses of the adjustment than same-spectrum noise. In Caswell-Midwinter and Whitmer (2020), it was further speculated that the large preference thresholds were due in part to the short duration of the stimuli. The current study tested this by measuring preference thresholds for gain adjustments across various stimulus durations. Although patients typically make quick comparisons on adjustments in the clinic, audiologists may talk for longer, which might elicit more frequent and reliable preferences.

Previous psychophysical research provides some evidence that speaking longer would lead to more consistent preferences: level discrimination improves with increasing duration, albeit mostly limited to short pure-tone stimuli. Increasing the duration of a 0.25, 1 or 8-kHz tone up to 0.5, 1 or 2 s, respectively, can improve level discrimination for normal-hearing listeners (Florentine, 1986). Further, duration can improve pure-tone level discrimination in fixed and roving pedestal level but not across-frequency conditions (Oxenham and Buus, 2000). For the discrimination of a tone’s relative level within a complex (i.e., profile analysis), performance improves up to a duration of at least 100 ms (Green et al., 1984; Dai and Green, 1993). The ability to discriminate a gain adjustment in particular band(s) of speech bears partial resemblance to increment detection, the detection of an increase or ‘bump’ in the level of an ongoing sound. Valente et al. (2011) showed that increasing the duration of an ongoing 0.5 or 4.0-kHz tone increased the detectability of a time-centred bump in the tone’s level more so than increasing the duration of the bump. There is some evidence of a duration effect with broadband stimuli: studying the detection of an 8-dB peak at 3.5 kHz in a broadband noise, Farrar et al. (1987) found that thresholds decreased as duration increased up to 300 ms, the maximum duration tested. Isarangura et al. (2019) found that the detection of spectral modulation in a broadband noise carrier also improved with increasing duration but reached asymptote by 200 ms. For speech stimuli, measures of duration effects on level discrimination are scant; in a study of overall level discrimination of speech, the threshold for words (mean duration 450 ms) was only significantly worse (greater) than for sentences (mean duration 1533 ms) when participants were aided (Whitmer and Akeroyd, 2011).

In sound-quality evaluations such as comparing hearing-aid settings, a balance must be struck in sound-sample duration. The sample must be long enough to allow perception of the acoustic changes, but short enough to allow comparison of the adjusted sound with the previous (reference) sound. The International Telecommunication Union (ITU) recommendations for subjective sound-quality evaluations note that, for paired comparisons, durations should not exceed 15-20 s due to “short-term human memory limitations,” but can be “a few seconds” (ITU, 2019, p. 6; cf. Cowan, 1984). These memory limitations - the ability to maintain features of the first sound for comparison to the second - are often measured by assessing the effect of the inter-stimulus interval (ISI) behaviourally (Winkler and Cowan, 2005) or physiologically (Bartha-Doering et al., 2015). In the clinic, the adjustment is often done without any gaps other than the natural pauses in ongoing speech. The memory limitation for comparing ongoing stimuli has previously been modelled as an exponential decay over many seconds, albeit for pure-tone stimuli (Durlach and Braida, 1969; Massaro, 1970). Despite qualitative recommendations and a long history of auditory memory research (cf. Cowan, 1984), the effect of duration on preferences for speech stimuli, as assessed in the clinic during hearing-aid adjustments, is not known.

On the basis of the foregoing evidence, we hypothesized that increasing the duration of the stimuli would elicit more consistent and reliable preferences for gain adjustments. The current study used most of the same methods, including most of the same participants, as Caswell-Midwinter and Whitmer (2020) did when measuring preferences for gain adjustments. The main difference is the primary experimental contrast: stimulus duration. To avoid potential memory confounds, the maximum stimulus duration was 6 s (cf. ITU-R 2003); the minimum was 2 s (vs. 0.855-2.3 s in the previous study). To better mimic elements of a clinical session, there were five other methodological differences. First, the stimuli were consecutive segments from a continuous story instead of repeated (within a trial) sentences. Second, the gain adjustment was always made for the second stimulus on each trial, rather than randomised. Third, the number of gain steps was reduced from six (±4, 8 and 12 dB) to four (±6 and 12 dB). Fourth, there was no ISI. Finally, given the lack of agreement or reliability in using descriptors (e.g., “tinny”) to describe the effect of a gain adjustment reported by Caswell-Midwinter and Whitmer (2020), the current study only measured preferences.

## Methods

### Participants

Twenty-nine adults (14 female) were recruited from a sample who had previously participated in a gain-discrimination experiment (Caswell-Midwinter and Whitmer, 2019). The median age was 68 years (range 51-74 years). The median better-ear four-frequency (0.5, 1, 2 and 4 kHz) pure-tone threshold average (BE4FA) was 35 dB HL (range 12-56 dB HL; see left panel of Figure 1). None of the participants had a conductive loss (i.e., all participants’ average air-bone threshold differences were less than 20 dB; British Academy of Audiology, 2016).

**Figure 1.**
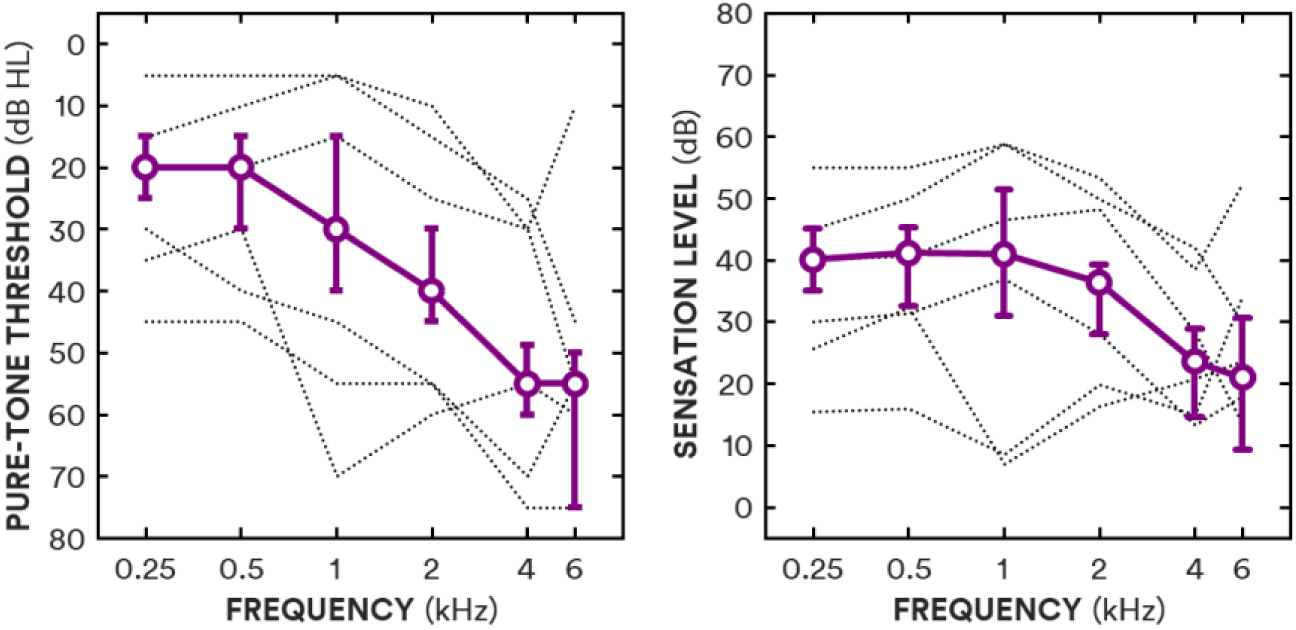
The left panel shows median pure-tone thresholds as a function of frequency (circles, solid line) and interquartile ranges (error bars), with the individual thresholds for the three lowest and highest average thresholds (dotted lines). The right panel shows median sensation levels (approximated from pure-tone thresholds and applied gain) as a function of frequency (circles, solid line) and interquartile ranges (error bars), with the individual values for the three lowest and highest average sensation levels (dotted lines).

For the 19 participants who habitually wore hearing aids at the time of the study, the real-ear insertion gain provided by their hearing aids in their better ear was measured with 65 dB broadband noise input (ICRA URGN-M-N; Dreschler et al., 2001) and used as their gain prescription. For the ten participants who were not currently wearing hearing aids, linear NAL-R gain prescriptions (Byrne and Dillon, 1986) for their better ear were used. Sensation level (SL) of the stimuli was approximated from pure-tone thresholds and applied gain; the median sensation level for amplified stimuli, averaged across 0.5, 1, 2 and 4 kHz, was 35 dB SL (range 15-51 dB SL; see right panel of Figure 1). All participants had previously been fit with hearing aids; the median hearing-aid experience was 10 years (range 2-35 years). Twenty-six of the 29 participants took part 18 months earlier in the preference experiment with short sentences (Caswell-Midwinter and Whitmer, 2020).

All participants had also performed visual letter and digit monitoring tasks during a previous study (at least 18 months prior to the current study) to provide an estimate of their cognitive abilities (specifically working memory; Gatehouse et al., 2006). The tasks involved identifying triplet digit and letter sequences at two different ISIs (1 and 2 s); a full description is in Caswell-Midwinter and Whitmer (2019b). The resulting *d’* measures were averaged across digit and letter tasks and ISIs to give a single cognitive score.

### Stimuli

The stimuli were consecutive segments of a Sherlock Holmes story read by a professional male actor with a Southern English accent (“The Naval Treaty”; Doyle, 2011). The original stimuli were converted from stereo to mono and resampled to 24 kHz from an original sample rate of 44.1 kHz. Any silent gaps greater than 250 ms were truncated to 250 ms. On each trial, two consecutive segments were presented to the participants’ better ear, both with the same duration of either 2, 4 or 6 s. For each segment, 50-ms linear onset and offset ramps were applied. To better mimic adjustments in the clinic, the standard stimulus was always the first stimulus in the pair, and there was no ISI beyond the offset and onset gating.

For the standard stimulus, real-ear or prescribed gain was applied across six frequency bands: a low-pass band with an upper cutoff of 0.25 kHz, four octave bands centred at 0.5, 1, 2 and 4 kHz, and a high-pass band with a lower cutoff of 6 kHz. For the target stimulus, additional gain (ΔGain) of either -12, -6, 0, +6 and +12 dB was applied in one of three broad frequency bands: a low-frequency band combining 0.25 (low-pass) and 0.5 kHz (octave) bands (LF), a mid-frequency band combining 1 and 2 kHz octave bands (MF), and a high-frequency band combining the 4 kHz and 6 kHz (high-pass) bands (HF). Stimuli were generated by convolving each segment with a 140-tap finite impulse response filter optimised for NAL-R equalisation at 24-kHz sample rate by Kates and Arehart (2010). The overall long-term A-weighted presentation level was 60 dB SPL to approximate in-quiet conversational level (Olsen, 1998). The presentation level was verified with an artificial ear and sound level meter (Brüel & Kjær 4152 and 2260), prior to any prescription or gain adjustment. The audibility of the segments was confirmed with each participant after the first trial.

We additionally analysed the effect of the natural variation in power within bands across the consecutive segments of each trial (i.e., when ΔGain = 0). There were significant mean absolute level differences within bands between the two segments in any given trial as a function of both frequency band and segment duration [F(2,56) = 13.06 and 19.41, respectively]. The differences, however, were small; absolute differences in band-specific level increased from 0.2 dB for the LF band to 0.3 dB for MF and HF bands [*t*(28) = 4.76; *p* ≪ 0.001], and absolute level differences decreased from 0.3 to 0.2 to 0.1 dB when the duration increased from 2 to 4 to 6 s, respectively [*t*(28) = -2.58 and -4.39; *p* = 0.015 and 0.0002, respectively].

### Procedure

Participants were seated in a sound-isolated booth (IAC Acoustics), and listened to the stimuli through circumaural headphones (AKG K702) without hearing aids. The change in stimulus within each trial from first to second segment was indicated on a touch screen in front of the participant. Participants were asked on each trial to indicate “How did the second sound compare to the first sound?” by selecting either the “better”, “worse” or “no difference” button on the touch screen.

There were three segment durations (2, 4 and 6 s) and 13 gain adjustments (±6 and ±12 dB adjustments in the LF, MF and HF bands plus a no-adjustment control), resulting in 39 stimulus conditions. Each stimulus condition was repeated ten times, resulting in 390 trials (3×13×10). The order of presentation was randomised for each participant. The trial run was broken into equal blocks of 130 trials with breaks between. Prior to testing, each participant completed 12 practice trials consisting of one trial each of 2-s and 6-s segments with ±12 dB gain adjustments in each of the three bands.

Ethical approval for the study was given by the West of Scotland research ethics committee (18/WS/0007) and NHS Scotland R&D (GN18EN094). All participants provided written informed consent prior to testing.

## Results

### Preferences

The proportions of “better” (B), “worse” (W) and “no difference” (ND) judgments were calculated for each gain adjustment in each frequency band (see Figure 2). A repeated-measures analysis of variance (RMANOVA) was run on the entire dataset (5 gain adjustments × 3 frequency bands × 3 segment durations) using combined “better” and “worse” proportions [P(B or W)] as the dependent variable (see Table 1). Amount of gain adjustment, frequency band and duration all showed significant main effects on better-and-worse preferences. Better and worse judgments increased with increasing duration, from 2 to 4 s [*t*_(28)_ = 8.44; *p* ≪ 0.001] and 4 to 6 s [*t*_(28)_ = 2.80; *p* = 0.0092]. The greatest rates of “better” and “worse” responses were for LF adjustments.

**Table 1.** Results of a repeated-measures analysis of variance on proportions of preferences, showing degrees of freedom (*df*), *F*-statistics and *p* values and partial eta-squared effect sizes. Degrees of freedom (*df*) and probabilities (*p*) reflect Greenhouse-Geisser (1959) corrections for non-sphericity.

| Main effects | $df$ (effect, error) | $F$ | $p$ | $\eta^2$ |
| --- | --- | --- | --- | --- |
| Band | 1.46, 40.81 | 128.30 | $<< 0.001$ | 0.82 |
| Gain | 2.92, 81.72 | 376.12 | $<< 0.001$ | 0.93 |
| Duration | 1.86, 52.05 | 55.20 | $<< 0.001$ | 0.66 |
| Interactions |  |  |  |  |
| Band · Gain | 5.10, 142.68 | 43.24 | $<< 0.001$ | 0.61 |
| Band · Duration | 3.76, 105.30 | 2.14 | 0.085 | 0.07 |
| Gain · Duration | 5.64, 157.88 | 4.87 | 0.0002 | 0.15 |
| Band · Gain · Duration | 8.05, 225.30 | 2.28 | 0.023 | 0.08 |

**Figure 2.**
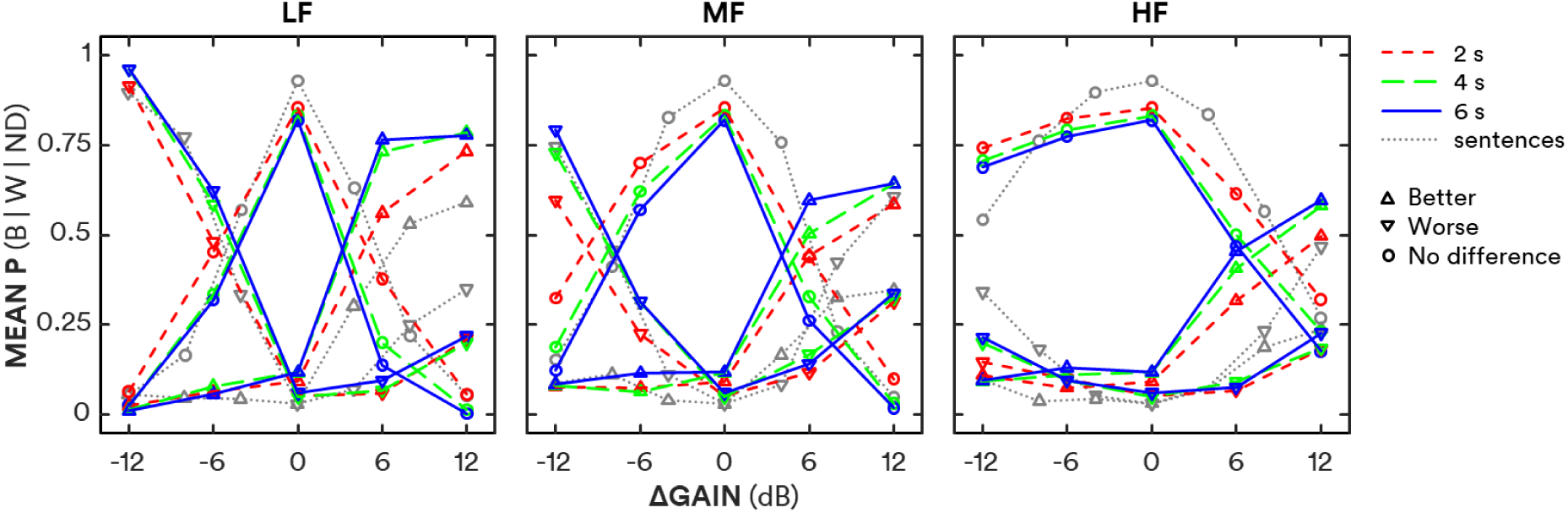
Mean proportion of preferences as a function of gain adjustment for low-frequency (LF; ≤ 0.5 kHz), mid-frequency (MF; 1-2 kHz) and high-frequency (HF; ≥ 4 kHz) bands (left, middle and right panels, respectively) for 2-s, 4-s and 6-s durations (red short-dashed, green long-dashed and blue solid lines, respectively). Better, worse and no difference preferences are shown as upward triangles, downward triangles and circles, respectively. Grey dotted lines and symbols show results using short sentences from Caswell-Midwinter and Whitmer (2020).

As the current methods shared many aspects, including participants, with Caswell-Midwinter and Whitmer (2020), the current study’s preference data were compared to the preferences elicited for short sentences in that previous study (grey triangles and dotted lines in Figure 2). In the current study there were more “better” and less “worse” ratings for +12-dB adjustments in the MF band [*t*(59) = 3.11 and -3.10 for better and worse, respectively; Holm-Bonferroni corrected *p*’ = 0.0028 and 0.0030] and HF band [*t*(59) = 5.32 and -3.77, respectively; both *p’ <* 0.001], There were also more “better” and less “worse” ratings for the LF band for +12 dB adjustments in the current study compared to the previous (compare grey with coloured triangles in the left panel of Figure 2), but these differences were not statistically significant [*t*_(59)_ = 1.99 and -1.60; both p > 0.05].

Participants were less prone to choose “no difference” when there was no gain adjustment in the current study compared to the previous study. The proportion of no difference responses at ΔGain = 0 was 0.84 across segment durations compared to 0.94 previously for short sentences [t(s6) = 3.31; p = 0.0017].

### Preference thresholds

The minimum gain adjustment required to elicit either a “better” or “worse” preference - the preference threshold - was estimated by fitting a logistic function to each individual’s P(B or W) as a function of ΔGain. Separate functions were fitted for negative and positive gain adjustments (i.e., decrements and increments) for each frequency band. The threshold was defined as P(B or W) = 0.55 [P(ND) = 0.45] which corresponds to *d*’ = 1 for an unbiased differencing observer in a same-different discrimination task (Macmillan and Creelman, 2005). Shapiro-Wilk tests of normality were violated for three of the 18 conditions: 4-s and 6-s LF increment and 2-s MF decrement thresholds (*W =* 0.91, 0.87 and 0.88, respectively; *p* = 0.018, 0.0034 and 0.0064); nevertheless, Tukey boxplots (Tukey, 1977) are used in Figure 3 to show the range of preference thresholds for each condition. All statistical probabilities reported for pairwise comparisons and correlations were corrected for multiple comparisons using the Holm-Bonferroni method (Holm, 1979); corrected probabilities are indicated by *p* ’.

**Figure 3.**
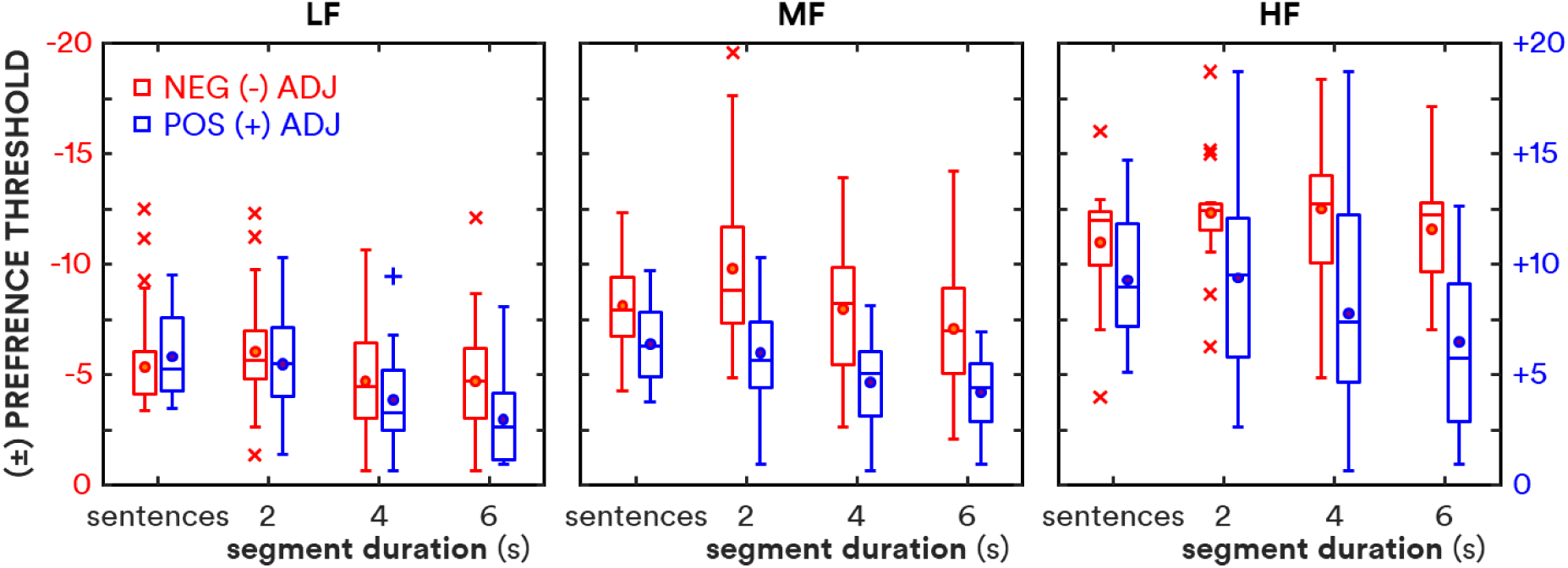
Boxplots of preference thresholds for each stimulus duration: sentences (average duration 1.6 s; Caswell-Midwinter and Whitmer, 2020), 2 s, 4 s and 6 s. Preference thresholds for negative and positive gain adjustments are shown in red and blue, respectively. Circles show means; lines show medians; boxes show interquartile ranges (IQR); whiskers show 1.5-IQR; crosses and pluses show outliers for negative and positive adjustments, respectively.

An RMANOVA based on the preference thresholds showed main effects of frequency band, direction of gain adjustment and segment duration (see Table 2). Preference thresholds decreased with increasing segment duration, increased with increasing centre frequency and were greater for decrements than increments. There was a significant interaction of frequency band and gain direction; decrement thresholds increased more than increment thresholds with increasing centre frequency. There was also a significant albeit modest (η^2^ = 0.11) interaction between gain direction and duration; preference thresholds decreased with increasing duration more for increments than decrements. There was additionally a significant but modest three-way interaction in the RMANOVA: preference thresholds for the MF band decreased with increasing segment duration more for decrements than for increments.

**Table 2.** Results of a repeated-measures analysis of variance on preference thresholds (see Table 1 for description of terms).

| Main effects | *df* (effect, error) | *F* | *p* | $\eta^2$ |
| --- | --- | --- | --- | --- |
| *Band* | 1.65, 46.34 | 139.05 | $\ll 0.001$ | 0.83 |
| *Direction* | 1, 28 | 70.80 | $\ll 0.001$ | 0.72 |
| *Duration* | 1.91, 53.38 | 48.43 | $\ll 0.001$ | 0.63 |
| Interactions |  |  |  |  |
| *Band · Direction* | 1.69, 47.33 | 11.54 | $\ll 0.001$ | 0.29 |
| *Band · Duration* | 2.94, 82.27 | 1.24 | 0.30 | 0.04 |
| *Direction · Duration* | 1.66, 46.49 | 3.35 | 0.042 | 0.11 |
| *Band · Direction · Duration* | 3.52, 98.69 | 3.76 | 0.0066 | 0.12 |

Mean thresholds with 95% repeated-measures confidence intervals (Loftus and Masson, 1994) are shown in Table 3. Thresholds significantly decreased with increasing duration for gain increments in the LF, MF and HF frequency bands, and for gain decrements in the LF and MF bands; the thresholds for decrements in the HF band (12.1 dB) did not significantly change across durations. The overall rate of change in preference threshold (i.e., the difference in mean thresholds not including HF decrements divided by the difference in duration) decreased with increasing duration from -0.8 dB/s at 4 s to -0.4 dB/s at 6 s. That is, preference thresholds decreased more between 2 and 4 s than between 4 and 6 s.

**Table 3.** Mean preference thresholds (dB) with 95% confidence intervals in brackets for all conditions (“-” = decrements; “+” = increments) including mean data from Caswell-Midwinter and Whitmer (2020), denoted “sentences.”

|  | LF |  | MF |  | HF |  |
| --- | --- | --- | --- | --- | --- | --- |
|  | - | + | - | + | - | + |
| **sentences** | 5.3<br>[4.6-6.1] | 5.8<br>[5.1-6.6] | 8.1<br>[7.4-8.9] | 6.4<br>[5.6-7.1] | 11.0<br>[10.2-11.8] | 9.3<br>[8.5-10.0] |
| **2 s** | 6.0<br>[5.3-6.8] | 5.5<br>[4.7-6.2] | 9.8<br>[9.0-10.6] | 6.0<br>[5.2-6.8] | 12.3<br>[11.6-13.1] | 9.4<br>[8.6-10.1] |
| **4 s** | 4.7<br>[4.0-5.5] | 3.9<br>[3.1-4.6] | 8.0<br>[7.2-8.7] | 4.7<br>[3.9-5.4] | 12.5<br>[11.7-13.3] | 7.8<br>[7.0-8.5] |
| **6 s** | 4.7<br>[3.9-5.5] | 3.0<br>[2.2-3.7] | 7.1<br>[6.3-7.9] | 4.2<br>[3.4-5.0] | 11.6<br>[10.8-12.3] | 6.5<br>[5.7-7.2] |

The preference thresholds measured here for 2-s consecutive segments of a continuous story were similar to the thresholds for short sentences reported by Caswell- Midwinter and Whitmer (2020) with the exception of MF and HF decrements, for which the current thresholds were significantly greater (*t* = 2.75 and 2.49; *p’ =* 0.011 and 0.030, respectively). Thresholds for 2-s stimuli, averaged across frequency bands, were positively correlated with thresholds in the previous study for both increments and decrements (ρ = 0.55 and 0.72, respectively; both *p’*≪ 0.001). Preference thresholds were not correlated with age, BE4FA, or hearing-aid experience (all *p’ >* 0.05). HF increment preference thresholds were positively correlated with HF pure-tone thresholds (ρ - 0.48; *p’ =* 0.049), and negatively correlated with HF sensation level (ρ = -0.50; *p’ =* 0.038) and cognitive score (*r =* -0.62; *p*’= 0.0020). Individual 2-s preference thresholds were correlated with individual decreases in threshold with duration, characterised as the slope in dB/s (*r* = -0.57; *p’=* 0.0035). Individual 2-s, 4-s or 6-s preference thresholds were not correlated with individual cognitive scores (*r =* -0.37, -0.13 and 0.03, respectively; all *p’* > 0.05), but slopes (dB/s) were correlated with cognitive scores (r = 0.50; *p =* 0.0057). Controlling for the variance shared with 2-s thresholds, individual slopes were still correlated with cognitive scores (r = 0.38; *p =* 0.047). That is, thresholds decreased more with duration (i.e., greater negative slope) for those with lesser letter/digit-monitoring ability. Based on this correlation, the RMANOVA of preference thresholds was re-run with centred cognitive scores as a covariate. As expected, the covariate reduced the error term, increasing the *F* statistics and η^2^ effect sizes, but did not change the pattern of results shown in Table 2.

### Preference agreement and reliability

Fleiss’ κ (Fleiss, 1971) was used to measure inter-participant agreement, comparing participants’ most frequent judgment (better, worse or no different) for each adjustment condition. To simplify the analysis, judgments were collapsed across adjustments for each direction and frequency band; judgments for the ΔGain = 0 condition were not included in the analysis. Fleiss’ κ was 0.39 [0.36-0.42 95% confidence intervals (CI)], 0.50 (0.47-0.53) and 0.50 (0.47-0.53) for segments of 2-s, 4-s and 6-s duration, respectively, representing “fair” (2 s) and “moderate” (4 and 6 s) agreement. That is, agreement significantly increased from 2-4 s, but not from 4-6 s.

A participant’s judgments (“better,” “worse” or “no difference”) for a given gain adjustment in a given frequency band were considered reliable if seven or more of those judgments were identical, a reliability threshold based on binomial probability theory (Kuk and Lau, 1995). Individual reliabilities were averaged across conditions; judgments for the ΔGain = 0 condition were not included. Because the proportions of reliable preferences in the current study were not normally distributed based on Shapiro-Wilk tests (*W* = 0.92, 0.90 and 0.92 for 2-s, 4-s and 6-s stimuli), non-parametric tests were used to compare reliability across conditions. Figure 4 shows individual proportions of adjustments with reliable preferences. Reliability increased significantly from a median value of 67% for short sentences and 2-s segments to 75% for 4-s and 6-s segments [*χ*^2^ = 11.10; *p* = 0.011]. There was no significant difference in reliability between sentences and 2-s segments (*z* = 0.65; *p* = 0.51) nor between 4-s and 6-s segments (*z* = 0.72; *p* = 0.47). The percentage of participants with ≥ 90% reliable preferences, however, did increase from 14% at 4 s to 28% at 6 s. Individual reliabilities for short sentences and 2-s stimuli were not correlated, but reliabilities for 4-s and 6-s stimuli were (*r* = 0.61; *p* = 0.0004).

**Figure 4.**
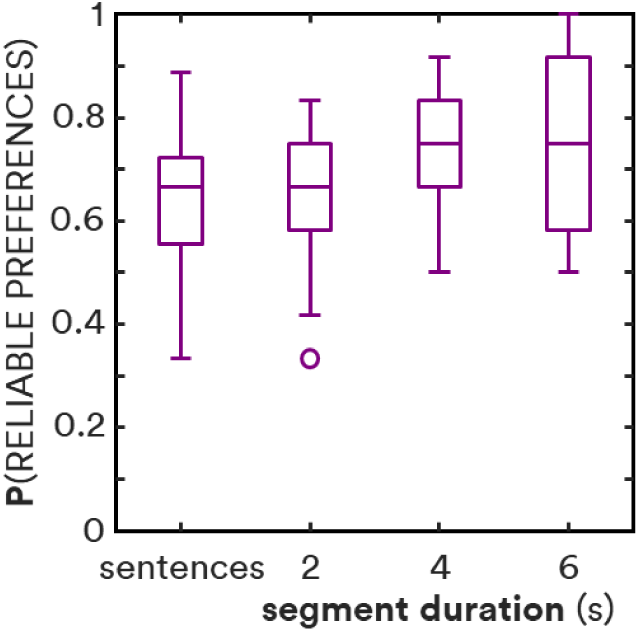
Proportion of reliable preferences as a function of stimulus duration. Horizontal lines show medians; boxes show interquartile ranges (IQR); whiskers show 1.5 • IQR; circles show outliers. Sentence data are from Caswell-Midwinter and Whitmer (2020).

## Discussion

By having participants compare and judge consecutive segments of a single-narrator story, we have shown that longer durations promote more frequent and reliable “better” or “worse” preference judgments for gain adjustments in broad frequency bands. That is, the gain adjustments required to elicit consistent preferences decreased with increasing stimulus duration. The proportions of better or worse preferences were greater, so preference thresholds were smaller, for increments than for decrements, in agreement with Caswell-Midwinter and Whitmer (2020) as well as previous psychophysical literature (Ellermeier 1996; Moore et al. 1989; Moore et al. 1997). Better and worse preferences were less frequent with increasing centre frequency of the adjustment band, as previously shown for short sentences (Caswell-Midwinter and Whitmer, 2020).

Despite differences in the method, the median preference thresholds in the current study for 2-s segments were similar to the thresholds for 1.6-s average duration sentences in our previous study (Caswell-Midwinter and Whitmer, 2020), and individual preference thresholds were correlated with the previous thresholds. As with the previous study, the strongest preferences were for increased LF gain and against decreased LF gain, as found in self-fitting studies (Keidser and Convery, 2018; Nelson et al., 2018; Vaisberg et al., 2021). The long-term spectrum of the stimuli had its greatest power in the LF band; this may have influenced the discriminability of LF adjustments (Jesteadt et al. 2017), increasing preferences and reliability. There were preference differences between the two studies, with increases in “better” vs. “worse” judgments for MF and HF increments in the current study. The long-term spectrum in the HF band for the current monologue segments was 5.6 dB less than for the previous sentence stimuli. Increases in HF gain may have then been judged more favourably in the current study because of the greater audibility in that band. There were, though, no spectral differences to explain the MF increment preference discrepancy; further work is needed to better understand to what extent particular stimulus attributes (e.g., vocal timbre) and context (e.g., monologue vs. unconnected sentences) affect gain preferences.

Participants were less likely to respond “no difference” in the current study when consecutive segments were presented without gain adjustments compared to the previous study (Caswell-Midwinter and Whitmer, 2020) where the same sentence was presented twice on each trial. This difference can be attributed to the comparison of two different speech segments; the naturally occurring differences in the spectrotemporal patterns between the two segments (without gain adjustments) could decrease the likelihood of a “no difference” response (Mason et al., 1984; Kidd et al., 1986). The effect of this decrease in no-difference responses on threshold estimation was minimal; fitting logistic functions to the current data using the no-difference responses from the previous study increased threshold estimates by only 0.4 dB on average. Nevertheless, the change demonstrates a limitation of using sequential stimuli for comparison.

The use of an ongoing story, as opposed to hearing the same utterance twice, anecdotally provided a greater degree of participant engagement with the material, engagement as might occur in the clinic, where the responses of the patient will affect real-world use. Any greater engagement with the stimulus content, however, may have been detrimental to performing the task. Beyond the decrease in no-difference responses, the effect of comparing different stimuli (two consecutive segments) versus comparing identical stimuli was small. Using non-repeating segments introduces variability in the level and spectrum in the comparison, which can decrease detectability (Kidd et al. 1986), thus increasing preference thresholds. In the present experiment, the use of the same talker throughout would have reduced signal uncertainty and thus reduced any effect of non-repeating segments on thresholds. To check the potential influence of extreme spectral variations between segment pairs, preference thresholds were recalculated excluding the 10% of trials with the greatest absolute difference in any band for each participant. The only significant effects of this recalculation were modest increases in the preference thresholds for 6-s MF and 2-s HF increment stimuli (Δthreshold = 0.2 and 0.3 dB; *z* = 2.72 and 2.13; *p* = 0.0065 and 0.032, respectively); all other threshold differences were not significantly different from zero (*z* = 0.14-1.22; all *p* > 0.05). Further, excluding trials based on extreme variation between their consecutive segments did not have any effect on the rate of change of preference thresholds as a function of duration. Thus, there is scant evidence that the natural variation in the consecutive stimuli affected the pattern of results.

The delivery of stimuli used for appraisal by the patient in the clinic may be different to paired or sequential comparisons. Rather, the appraisal may take the form of a single interval. Single interval ratings of hearing-aid sound quality have shown moderate test-retest reliability (Narendran and Humes, 2003) and good inter-rater reliability (Gabrielsson et al. 1990), but these studies used stimulus durations of 50-60 s. Using such long stimuli for clinical fine-tuning may not be feasible.

It is not known if durations > 6 s would provide even greater discriminability and more reliable preferences. While the thresholds across most conditions decreased significantly from 4 s to 6 s, the effect was small. The overall rate of change decreased from -0.8 dB/s between 2 and 4 s to -0.4 dB/s at 6 s, resembling the exponential decay in memory-based models of the effects of duration on pairwise comparison (e.g., Durlach and Braida, 1969). There was a correlation between participants’ monitoring-task cognitive scores and the rate of decrease in their preference thresholds with increasing duration. That is, the worse their cognitive scores, the stronger the effect of stimulus duration on preference thresholds. This suggests that there is a limit to the effect of duration in the judgment of gain adjustments, and further suggests that the greatest effect is for those with lesser cognitive capacity. The mean preferences were very similar for 4-s and 6-s stimuli (Figure 2), and there was no increase from 4 to 6 s in inter-participant agreement or intra-participant reliability (Figure 4). It is therefore unlikely for thresholds to decrease, or reliability to increase, much further beyond the results here for 6-s stimuli (cf. Bartha-Doering et al., 2015). It is also not known how fast-acting compression, as delivered by many current hearing aids, would affect results. The short-term variation in speech would interact with the compressor, potentially generating different preferences. The dynamic compression of speech, however, has previously not been found to have an effect on overall level discrimination of words and sentences (Whitmer and Akeroyd, 2011), hence would not be expected to lead to more consistent preferences with duration.

The improvement in thresholds and reliability with increasing stimulus duration was small relative to the thresholds and reliabilities themselves. Talking or presenting stimuli for 6 s to a hearing-aid wearer in the clinic would help elicit consistent preferences for adjustments, but those adjustments would still need to be large: 3-6 dB for increments, 5-12 dB for decrements. These thresholds are well above common troubleshooting adjustments, especially for adjustments at higher frequencies. A patient may indeed state an immediate preference when a smaller adjustment has been made, but such a preference should be treated with caution, as it may not be based on the acoustical percept of the adjustment, and is therefore likely to be unreliable. For the personalisation of hearing aids in the clinic, it is therefore important not only to say more than a few words (e.g., “how’s that sound?”) immediately following an adjustment, but also to ensure that the adjustment is large enough to elicit a consistent effect. Given these constraints, alternative methods of fitting, such as self-adjustments (Boothroyd and Mackersie, 2017; Nelson et al., 2018), which have resulted in similar gains to those prescribed and fit by a clinician (cf. Sabin et al., 2020), may be more viable for effective hearing-aid personalisation, although further study is warranted.

## Data Availability

Data available upon request

## Acknowledgments

The authors would like to thank David McShefferty for his assistance in conducting the study, as well as Dr. Gitte Keidser, Prof. Brian C. J. Moore and two anonymous reviewers for their helpful comments. This work was supported by funding from the Medical Research Council [grant numbers MR/S003576/1 and 1601056]; and the Chief Scientist Office of the Scottish Government.

## References

Anderson MC, Arehart KH, Souza PE (2018) Survey of current practice in the fitting and fine-tuning of common signal-processing features in hearing aids for adults. J Am Acad Audiol 29:118–124. DOI: 10.3766/jaaa.16107.

Bartha-Doering L, Deuster D, Giordano V, am Zehnhoff-Dinnesen A, Dobel C (2015) A systematic review of the mismatch negativity as an index for auditory sensory memory: From basic research to clinical and developmental perspectives. DOI: 10.1111/psyp.l2459

Bentler RA, Niebuhr DP, Johnson TA, Flamme GA. (2003). Impact of digital labelling on outcome measures. Ear Hearing 24(3), 215–224. DOI: 10.1097/01.AUD.0000069228.46916.92

British Academy of Audiology (2016) Guidance for Audiologists: Onward Referral of Adults with Hearing Difficulty Directly Referred to Audiology Services. Retrieved from: https://www.baaudiology.org/app/uploads/2019/07/BAA_Guidance_for_Onward_Referral_of_Adults_with_Hearing_Difficulty_Directly_Referred_to_Audiology_2016_-_minor_amendments.pdf.

Caswell-Midwinter B, Whitmer WM (2019) Discrimination of gain increments in speech. Trends Hear 23. DOI: 10.1177/2331216519886684.

Caswell-Midwinter B, Whitmer WM (2019b) Discrimination of gain increments in speech-shaped noises. Trends Hear 23. DOI: 10.1177/2331216518820220.

Caswell-Midwinter B, Whitmer WM (2020) The perceptual limitations of troubleshooting hearing aids based on patients’ descriptions. Int J Audiol 60(6): 427–437. DOI: 10.1080/14992027.2020.1839679.

Cowan N (1984) On short and long auditory stores. Psych Bull 96(2): 341–370. DOI: 10.1037/0033-2909.96.2.341.

Dai H, Green DM (1993) Discrimination of spectral shape as a function of stimulus duration. J Acoust Soc Am 93(2): 957–965. DOI: 10.1121/1.405456.

Dawes P, Hopkins R, Munro KJ (2013) Placebo effects in hearing-aid trials are reliable. Int J Audiol 52(7): 472–477. DOI: 10.3109/14992027.2013.783718

Doyle AC (2011) The Memoirs of Sherlock Holmes (D. Jacobi, narr.) [Audiobook]. London: AudioGO Ltd.

Dreschler WA, Verschuure H, Ludvigsen C, Westermann S (2001) ICRA noises: Artificial noise signals with speech-like spectral and temporal properties for hearing instrument assessment. Audiol 40(3): 148–157.

Durlach NI, Braida LD (1969) Intensity perception. I. Preliminary theory of intensity resolution. J Acoust Soc Am 46(2): 373–383. DOI: 10.1121/1.1911699.

Ellermeier W (1996) Detectability of increments and decrements in spectral profiles. J Acoust Soc Am 99(5): 3119–3125. DOI: 10.1121/1.414797.

Farrar CL, Reed CM, Ito Y, Durlach NI, Delhorne LA, Zurek PM, Braida LM (1987) Spectral-shape discrimination I: Results from normal-hearing listeners for stationary broadband noises. J Acoust Soc Am 81(4): 1085–1092. DOI: 10.1121/1.394628.

Fleiss JL (1971) Measuring nominal scale agreement among many raters. Psych Bull 76(5): 378–382. DOI: 10.1037/h0031619.

Florentine M (1986) Level discrimination of tones as a function of duration. J Acoust Soc Am 79(3): 792–798. DOI: 10.1121/1.393469.

Gatehouse S, Naylor G, Elberling C (2006) Linear and nonlinear hearing aid fittings - 2. Patterns of candidature. Int J Audiol 45(3): 153–171. DOI: 10.1080/14992020500429484.

Green DM, Mason CR, Kidd G (1984) Profile analysis: Critical bands and duration. J Acoust Soc Am 75(4): 1163–1167. DOI: 10.1121/1.390765.

Greenhouse SW, Geisser S (1959) On methods in the analysis of profile data. Psychometrika 24: 95–112. DOI: 10.1007/BF02289823.

Holm S (1979) A simple sequentially rejective multiple test procedure. Scand J Stat 6: 65–70.

International Telecommunication Union, Radiocommunication Sector (2019) General methods for the subjective assessment of sound quality. Recommendation ITU-R BS.1284-2

International Telecommunication Union, Telecommunication Standardization Sector (2003) Subjective test methodology for evaluating speech communication systems that include noise suppression algorithm. Recommendation ITU-T P.835.

Isarangura S, Eddins AC, Ozmeral EJ, Eddins DA (2019) The effects of duration and level on spectral modulation perception. J Speech Lang Hear Res, 62: 3876–3886.

Jenstad LM, Van Tasell DJ, and Ewert C (2003) Hearing Aid Troubleshooting Based on Patients’ Descriptions. J Am Acad Audiol, 14 (7): 347–360.

Jesteadt W, Walker SM, Oluwaseye AO, Ohlrich B, Brunette KE, Wróblewski M, Schmid KK (2017) Relative contributions of specific frequency bands to the loudness of broadband sounds. J Acoust Soc Am, 142(3): 1597–1610. DOI: 10.1121/1.5003778.

Kates JM, Arehart KH. (2010). The Hearing-Aid Speech Quality Index (HASQI). J Audio Eng Soc, 58: 363–381.

Keidser G, Convery E (2018). Outcomes with a self-fitting hearing aid. Trends Hear, 22: 1–12. DOI: 10.1177/2331216518768958.

Kidd G, Mason CR, Green DM (1986). Auditory profile analysis of irregular sound spectra. J Acoust Soc Am, 79(4): 1045–1053.

Kuk FK, Lau C (1995) The application of binomial probability theory to paired comparison responses. Am J Audiol, 4 (1): 37–42. DOI 10.1044/1059-0889.0401.37

Kuk FK, Ludvigsen C (1999) Variables affecting the use of prescriptive formulae to fit modern nonlinear hearing aids. J Am Acad Audiol, 10: 453–465.

Loftus GR, Masson MEJ (1994) Using confidence intervals in within-subject designs. Psychon Bull Rev, 1: 476–490. DOI: 10.3758/BF03210951

Macmillan NA, Creelman CD (2005) Detection theory: A user’s guide (2nd ed.). Mahwah, NJ: Lawrence Erlbaum Associates.

Mackersie CL, Boothroyd A, Lithgow A. (2019) A “Goldilocks” approach to hearing aid self-fitting: ear-canal output and speech intelligibility index. Ear Hearing 40(1): 107–115. DOI: 10.1097/AUD.0000000000000617.

Mason CR, Kidd G, Hanna TE, Green DM (1984) Profile analysis and level variation. Hear Res 13(3): 269–275. DOI: 10.1016/0378-5955(84)90080-7.

Moore BCJ, Oldfield SR, Dooley GJ (1989) Detection and discrimination of spectral peaks and notches at 1 and 8 kHz. J Acoust Soc Am 85: 820–836. DOI: 10.1121/1.397554.

Moore BCJ, Peters RW (1997) Detection of increments and decrements in sinusoids as a function of frequency, increment, and decrement duration and pedestal duration. J Acoust Soc Am 102(5): 2954–2965. DOI: 10.1121/1.420350.

Narendran MM, Humes LE (2003) Reliability and validity of judgments of sound quality in elderly hearing aid wearers. Ear Hear 24(1): 4–11. DOI: 10.1097/01.AUD.0000051745.69182.14.

Naylor G, Öberg M, Wänström G, Lunner T (2015) Exploring the effects of the narrative embodied in the hearing-aid fitting process on treatment outcomes. Ear Hear 36(5): 517–526. DOI: 10.1097/AUD.0000000000000157.

Nelson PB, Perry TT, Gregan M, Van Tasell D (2018) Self-adjusted amplification parameters produce large between-subject variability and preserve speech intelligibility. Trends Hear, 22. DOI: 10.1177/2331216518798264.

Oxenham AJ, Buus S (2000) Level discrimination of sinusoids as a function of duration and level for fixed-level, roving-level, and across-frequency conditions. J Acoust Soc Am 107(3): 1605–1614. DOI: 10.1121/1.428445.

Pollack I (1972) Memory for auditory waveform. J Acoust Soc Am 52(4): 1209–1215. DOI: 10.1121/1.1913234.

Sabin AT, van Tassell DJ, Rabinowitz B, Dhar S (2020) Validation of a self-fitting method for over-the-counter hearing aids. Trends Hear 24: 2331216519900589. doi: 10.1177/2331216519900589.

Thielemans TD, Pans M. Chenault, and Anteunis L (2017) Hearing aid fine-tuning based on Dutch descriptions. Int J Audiol, 56 (7): 507–515. DOL :10.1080/14992027.2017.1288302.

Vaisberg JM, Beaulac S, Glista D, Macpherson EA, Scollie SD (2021) Perceived sound quality dimensions influencing frequency-gain shaping preferences for hearing aid-amplified speech and music. Trends Hear, 25. DOI: 10.1177/2331216521989900.

Valente DL, Patra H, Jesteadt W (2011) Relative effects of increment and pedestal duration on the detection of intensity increments. J Acoust Soc Am 129(4): 2095–2103. DOI: 10.1121/1.3557043.

Whitmer WM, Akeroyd MA (2011) Level discrimination of speech sounds by hearing-impaired individuals with and without hearing amplification. Ear Hear 32: 391–398. DOI:10.1097/AUD.0b013e318202b620.

Winkler I, Cowan N (2005) From sensory to long-term memory: Evidence from auditory memory reactivation studies. Exp Psych 52(1): 3–20. DOI: 10.1027/1618-3169.52.1.3.

